# Atypical gut microbial ecosystem from athletes with very high exercise capacity improves insulin sensitivity and muscle glycogen store in mice

**DOI:** 10.1101/2024.09.27.24314273

**Authors:** David Martin, Mathis Bonneau, Luz Orfila, Mathieu Horeau, Mathilde Hazon, Romain Demay, Emmanuelle Lecommandeur, Rufin Boumpoutou, Arthur Guillotel, Pierrick Guillemot, Mickael Croyal, Pierre Cressard, Chrystèle Cressard, Anne Cuzol, Valérie Monbet, Frédéric Derbré

**Author notes:** **Co-corresponding authors:** Frederic Derbré, PhD, Assistant professor, Laboratory “Movement, Sport and Health Sciences” – EA7470, University of Rennes 2 / ENS Rennes, 35170 Bruz, France, Mail, Valerie Monbet, PhD, Full professor, IRMAR - UMR CNRS 6625, University of Rennes, 35000 Rennes, France, Mail. The last co-authors equally contributed to this work (Valérie Monbet, Frédéric Derbré).

## Abstract

**Background:** The gut bacterial ecosystem plays a key role in the host’s energy metabolism, and potentially in the host’s exercise capacity, recognized as a powerful predictor of health status and risk of mortality.

**Objective:** To deepen our understanding of the gut bacterial ecosystem relationship with host’s exercise capacity and energy metabolism, we characterized the gut microbiota in a cohort of healthy humans with heterogeneous exercise capacities, and next determined the impact of fecal microbiota transplantation (FMT) from donors of this cohort on energy metabolism and exercise capacity of recipient mice.

**Design:** 50 male normo-weight participants (from inactive to elite endurance athletes) performed food frequency questionnaire (FFQ) and exercise tests to determine exercise capacity parameters (VO_2max_, fat oxidation, exercise energy expenditure). Metagenomic shotgun and metabolomic analyses were performed to characterize gut microbiota ecosystem and short-chain fatty acids (SCFAs) on human and mice fecal samples. Mice performed running exercise capacity tests and metabolic parameters were determined in skeletal muscle and plasma samples.

**Results:** While our data support that the bacterial ecosystem appears to be modestly altered between individuals with low to high exercise capacities, we report that gut bacterial α-diversity, density, and functional richness are significantly reduced in athletes with very high exercise capacity. By using FMT, we report that the engraftment of these atypical gut microbiota improves insulin sensitivity and muscle glycogen stores into transfected mice.

**Conclusion:** Our data highlight promising therapeutic perspectives in fecal transplantation from human donors selected based on exercise capacity parameters.

*What is already known on this topic:*

- Gut microbiota ecosystem directly affects exercise capacity and muscle energy metabolism.
- Athletes with high exercise capacity exhibit greater gut microbiota diversity and an over-representation of some bacterial species compared to inactive individuals, but current available data present major bias including the lack of consideration of dietary habits and body composition.

*What this study adds:*

- Exercise capacity is associated with atypical gut microbiota communities, independently of food habits and body composition.
- Atypical gut microbial ecosystem from athletes with very high exercise capacity are related to high fecal propionate content, but negatively associated with gut microbiota *α*-diversity, bacterial density and gut microbiota functional abundance.
- Depending on the donor’s exercise capacity, gut bacterial ecosystems affect insulin sensitivity, but not exercise capacity, of transfected mice. How this study might affect research, practice or policy: - These data open promising research perspectives: 1) to improve the management of the gut microbiota ecosystem of elite athletes and patients performing adapted physical activity for therapeutic purposes, and 2) to personalize FMT in patients treated for non-communicable diseases by including exercise capacity parameters in the clinical criteria for donor selection.

## INTRODUCTION

Exercise capacity is now considered as a powerful predictor of health status and risk of mortality [1,2]. It refers to the maximum amount of physical exertion that an individual can sustain over a prolonged period of time [3]. Exercise capacity can be assessed during exercise testing by measuring maximal oxygen consumption 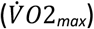 and fat oxidation (FO) through indirect calorimetry. 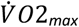 characterizes the maximum energy that an organism is able to produce during incremental exercise, while the percentage of FO during fasted submaximal exercise provides insight into the whole-body metabolic profile [4,5]. Specifically, during light to moderate submaximal exercise, a healthy organism preferentially degrades fatty acids for its energy supply, derived from both adipose tissue and intramuscular triglyceride breakdown [6]. When the intensity is high to maximal, the organism switches to preferentially use carbohydrates (CHO) provided by muscle glycogen and blood glucose derived from both liver glycogenolysis and gluconeogenesis [6]. However, unlike the almost unlimited fatty acid stores, muscle glycogen stores are limited, and their depletion leads to a reduction in exercise intensity or duration [7]. Increasing the proportion of energy substrates other than CHO during exercise, particularly fatty acids, is thus an essential strategy to spare muscle glycogen improving exercise capacity [8].

The gut bacterial ecosystem plays a key role in the host’s energy metabolism [9–11], and potentially in the host’s exercise capacity. In germ-free or antibiotics-treated (ATB) mice, the absence of gut microbiota directly reduces exercise capacity [12–14], notably due to concomitant reduction in intestinal absorption of glucose and muscle glycogen stores [13,15]. Moreover, the absence or reduction of short-chain fatty acids (SCFAs) production in an organism lacking gut microbiota or exhibiting dysbiotic gut microbiota displays the crucial role of SCFAs in these alterations [16,17]. In humans, cross-sectional studies reveal greater diversity and an over-representation of some bacterial populations in the gut microbiota of elite athletes compared to less active individuals [18–21]. However, all these clinical studies show some limitations including the lack of consideration of dietary habits and body mass index which are known to modulate the gut microbiota composition and function [22].

Herein, we report our characterization of the relationship between exercise capacity and the gut bacterial ecosystem in humans, ranging from inactive to elite endurance athletes, independently from diet habits or body composition ([EXOMIC cohort](https://clinicaltrials.gov/study/NCT05220657)). Our data highlight that exercise capacity impacts gut bacterial ecosystem and fecal SCFA levels, independently from dietary habits. Surprisingly, we observe that humans with very high exercise capacity exhibit reduced gut microbiota diversity, density and functional pathways abundance. Using fecal microbiota transplantation (FMT) from humans of our cohort to ATB-pretreated mice, we demonstrate that the gut microbiota associated to exercise capacity of donors affects insulin sensitivity and muscle glycogen store in transfected mice, highlighting the essential role of the gut microbiota associated to donor’s exercise capacity in the changes of metabolic profile of the FMT recipients.

## MATERIELS AND METHODS

### Human clinical study

The EXOMIC pilot clinical study took place at the laboratory “Movement, Sport and health Sciences” (M2S, Rennes, France). The study was approved by the national Comité de protection des personnes Ouest IV Nantes (ID-RCB: 2021-A02496-35) and registered on ClinicalTrials.gov under NCT05220657.

### Mouse experiments

C57BL/6J male mice (8-week-old; Janvier Labs, France) were randomly divided into the following two experimental groups: control group (CTL, n = 12) versus mice transfected (n = 24) by heterogeneous human donors (n=8). Except for running tests, mice were housed in individual standard cages without wheels, and thus were not subjected to daily exercise.

### Medical inclusion visits, fecal samples collection and exercise testing

Each participant completed the food frequency questionnaire to estimate daily food and nutrient intakes (FFQ) [23]. Daily energy expenditure was also calculated on a weekly basis (MET-minutes per week) using the Global Physical Activity Questionnaire (GPAQ) [24]. Percentage body fat (%FM) was measured by skinfold measurement using Harpenden® forceps based on the 4-skinfolds method. One fecal sample was collected for each participant during the 15 days following the medical inclusion and before the last visit to the laboratory. The participants performed two exercise tests on ergocycle to respectively determine (1) maximal oxygen consumption 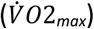 and (2) carbohydrate (CHO) and fat oxidation (FO) using an indirect calorimetry system (Ultima CardiO2, Medgraphics, United Kingdom).

### Fecal Material Transfection (FMT) from humans to mice

After 10 days of treatment with an oral cocktail of broad-spectrum antibiotics [13], mice selected for transfection (n=24) received 100 µg of fecal material by oral gavage every morning during 3 days, then once per week for the next two weeks. Fecal samples from FMT and CTL mice were individually collected before and after the antibiotic treatment, and 5 weeks post-FMT. Gastrocnemius muscles and abdominal fat mass were dissected, weighed and then either frozen in liquid nitrogen.

### Running exercise capacity in mice

All mice performed a submaximal running test on a treadmill (Ugo Basile, Gemonio, Italy) before and after antibiotic treatment, and then each week during the next 2 weeks. Running exercise capacity was determined by the time until exhaustion from a test adapted from Okamoto et al. [14].

### Metabolic parameters

Glycemia was measured using Automated Beckman Coulter (Beckman Coulter, Brea, CA). Serum insulin concentrations were measured by enzyme-linked immunosorbent assay (ELISA) according to manufacturer’s instructions (Millipore, St Louis, MO, USA). The acid-hydrolysis method used for muscle glycogen quantification was adapted from the protocols described by Adamo and Graham [25].

### Metagenomics data

Fecal metagenomic shotgun sequences from both human and mice experiments underwent a pre-processing pipeline, where sequences were quality filtered using KneadData2 with default parameters [26]. MetaPhlAn3 was used for quantitative profiling the taxonomic composition of the microbial communities of all metagenomic samples [26]. HUMANn3 was used to profile pathway and gene family abundances [26].

### Metabolomic data

Measurement of short-chain fatty acids (SCFAs), amino acids and bile acids (BAs) concentrations were determined in fecal samples by liquid chromatography-tandem mass spectrometry (LC-MS/MS) by the CORSAIRE platform (Biogenouest, Nantes, France).

### Statistical analysis

Data are presented as means ± SD. Statistical analysis between groups was analyzed according to the data distribution and data type. The threshold for statistical significance was assumed to be p<0.05. Additional experimental methods and information are provided in online supplemental materials files.

## RESULTS

### Exercise capacity is associated with atypical gut microbiota communities, independently of food and nutrient intakes

We first examined whether exercise capacity can affect gut microbiota compositions in healthy humans. To this purpose, we recruited 50 young males to create a physiological and biological database ranging from very low to very high exercise capacity populations, including non-athletes (NoA, n=21), elite soccer players (ESP, n=15) and elite cyclists (EC, n=14). To best limit the differences in food habits between experimental groups, we recruited the NoA participants by matching their food intakes collected by Short Form Food Frequency Questionnaire to correspond qualitatively to the EC’s food intakes (Figure 1A, Supplemental Material). All participants recruited were normo-weight (Table S1), completed a long form of FFQ, and performed two sessions of exercise testing (Figure 1A). Participants included in this cohort presented 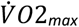 levels ranging from 35 to 85 ml/min/kg (Figure 1B), with a proportion of energy expenditure from fat oxidation during submaximal exercise, ranging from 0 to 85% (Figure 1C), characterizing a highly heterogeneous cohort in terms of energy expenditure and energy substrates used during exercise.

**Figure 1.**
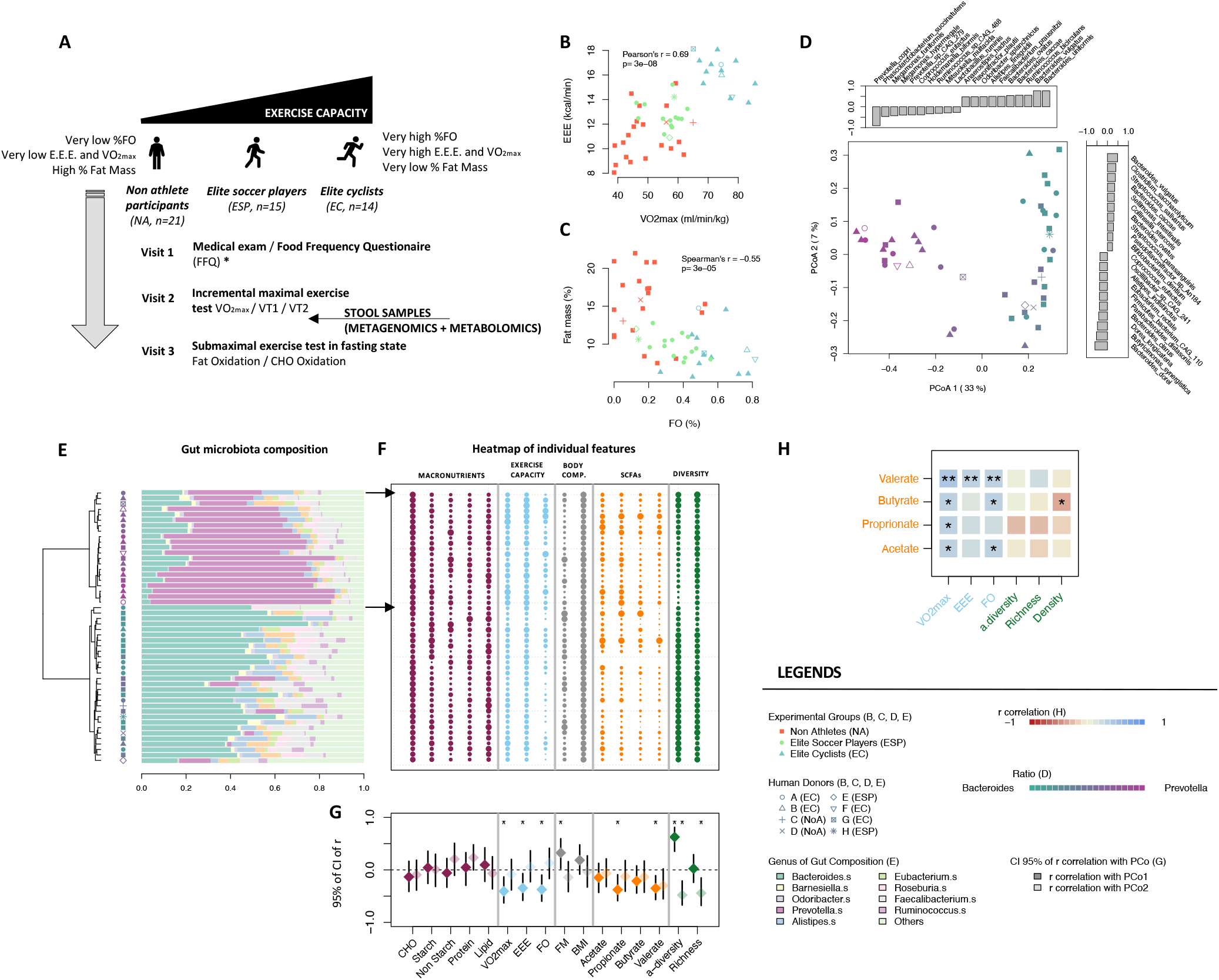
Exercise capacity is associated with gut microbiota communities, independently of food and nutrient intakes. (**A**) Experimental design (*non-athlete participants were selected to have their dietary food intake match that of elite athletes). (**B**) E.E.E (kcal/min) with respect to 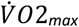 (ml/min/kg). (**C**) Fat mass with respect to Fat Oxidation (% of the total energy substrate oxidation). (**D**) Principal Coordinates Analysis (PCoA) from Bray Curtis dissimilarity of humans’ gut microbiota compositions. Subjects are colored by their ratio of *Bacteroides*/*Prevotella* within their metagenome. The bar plots represent the strongest Spearman’s r correlations (in absolute value) between species and PCoA coordinates. (**E**) Hierarchical Ascendant Clustering computed from the Bray Curtis pairwise dissimilarity matrix (left panel) with the gut microbiota composition of the participants (1 line = 1 participant; right panel) (**F**) Heatmap of relative macronutrient intakes, exercise capacity parameters, body composition, fecal SCFA content and gut microbiota composition’s diversities (1 line = 1 participant). The points’ width depends on the intensity of the variable (i.e. normalization by the intra-variable maximum is applied). (**G**) 95% confidence interval of the r correlation between each variable with PCo1 (in dark colors) and PCo2 (in light colors) (see panel **D**). (**H**) r correlation between exercise energy parameters, gut microbiota diversity and bacterial density with fecal SCFA content. Statistical significance of the correlation: **\*** p<0.05; **\*\*** p<0.01; **\*\*\*** p<0.001.

All participants provided their fecal samples to characterize the gut bacterial ecosystem using metagenomics sequencing [26]. We first observe that the two coordinates of the Principal Coordinates Analysis (PCoA) of the pairwise dissimilarity matrix of samples (i.e. Bray Curtis dissimilarity metric) account for 40% of the variability in gut microbiota composition within EXOMIC cohort (Figure 1D). Specifically, individuals with negative coordinates on PCo1 are predominantly elite cyclists and exhibit a gut microbiota composition characterized by a high relative abundance of species belonging to the *Prevotella* genus, mainly represented by the high expression of *Prevotella copri* (Figure 1D). Participants with positive coordinates on PCo 1 are mostly non-athletes and present a gut microbiota composition characterized by a high relative abundance of species belonging to the *Bacteroides* genus, associated with high expression of *Bacteroides uniformis, Bacteroides vulgatus* and *Feacalibacterium prausnitzii* (Figure 1D). These observations are consistent with previous findings [21,27,28], and support the concept that humans with high exercise capacity have an atypical gut microbiota composition compared to their counterparts with low exercise capacity.

On the Figures 1E and 1F, each line symbolizes variables characterizing a participant included in the EXOMIC cohort. The Hierarchical Ascendant Clustering (HAC) of gut microbiota compositions reveals two distinct clusters related to exercise capacity parameters (Figure 1E), close to the *Bacteroides* and *Prevotella* enterotypes concept [29]. The Figure 1F represents the environmental variables associated to each gut microbiota composition, ordered by the HAC. We first estimated the 95% of the confidence intervals for the r correlation coefficients between environmental variables (i.e. macronutrients, physiological variables, body composition, SCFAs and diversity indexes) and coordinates that summarize the variation of gut microbiota compositions within the EXOMIC cohort. Our results reveal that relative macronutrient intakes, including carbohydrates (CHO), fibers, proteins, and lipids, exhibit weak and non-significant association with the coordinates of the gut microbiota composition’s PCoA (Figures 1G and S1B). Employing non-parametric redundancy analysis (Canonical Analysis of Principal Coordinates, CAP, Supplemental Material), we measured how strongly environmental variables (i.e. diet, exercise capacity parameters, SCFAs) are linked to the structure and composition of the bacterial ecosystem, computing the explained variance and its associated p value. CAP confirms no discernible association between food or nutrient intakes and gut microbiota compositions (p>0.05, Figures S1C-F). Conversely, exercise capacity variables are significantly linked with 12% of the gut microbiota compositions (p=0.01, Figure S1G). Notably, 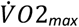, EEE and FO display positive and significant associations with the first Coordinate of the gut microbiota composition’s PCoA (p<0.05, Figure 1G). Together, these findings support that exercise capacity directly affects gut microbiota composition, independently of food or nutrient intakes.

### Atypical gut microbial ecosystem from athletes with very high exercise capacity are also related to high fecal propionate content

We report in our cohort a significant association of 15% between fecal SFCAs content and gut microbiota compositions (p=0.01, Figure S1H). Notably, fecal valerate and propionate content exhibit the strongest associations (p<0.05, Figure 1G) with the first Coordinate of the gut microbiota composition’s PCoA, mainly driven by the *Prevotella copri* expression (Figure 1D). Species belonging to the *Prevotella* genus are generally related to a non-western diet [30,31], where starch fermentation leads to an increase in fecal SCFA content [32]. Here, we observe that the proportion of starch intakes are not associated with fecal valerate and propionate content within the EXOMIC cohort (Figure S2A), suggesting that the host’s exercise capacity could impact fecal SCFA production independently of fiber intakes. Interestingly, *Phascolarctobacterium succinatutens*, well-identified to transform succinate into propionate [33], is the second main bacteria associated with the first Coordinate of the gut microbiota composition’s PCoA (Figure 1D). While *Prevotella copri* appears to be a succinate producer rather than propionate producer [34], it seems consistent that fecal propionate content and those species are associated with the first Coordinate of the gut microbiota composition’s PCoA. These associations between 1) high exercise capacity parameters, 2) gut microbiota composition dominated by *Prevotella copri* and *Phascolarctobacterium succinatutens* and 3) high fecal propionate content are finally consistent with previous studies supporting that propionate supplementation improves exercise capacity in mice [19], as well as resting fat oxidation and energy expenditure in fasted humans [35].

### High exercise capacity is negatively associated with gut microbiota *α*-diversity, bacterial density and gut microbiota functional abundance

Over the last decade, clinical studies conducted in humans with low to moderate exercise capacity have shown that 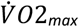 or training volume was positively correlated with gut microbiota *α*-diversity [28,36,37]. Here, our data support that gut microbiota *α*-diversity would ultimately follow an inverted-U shape relationship with 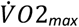 (Figure 2A). For hosts with 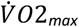 between 35 and 60 ml/min/kg, the association is positive and consistent with previous studies [36]. However, for hosts with 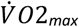 greater than 60 ml/min/kg, the association becomes negative (Figure 2A). Fecal bacterial density is also significantly reduced in hosts with a high capacity to consume energy during maximal exercise (Figure 2B). Interestingly, we also report a negative relationship between fat oxidation and gut microbiota *α*-diversity (p<0.05, Figure 2D), hosts with the lowest fecal bacterial density exhibiting the significant highest fat oxidation during submaximal exercise (p<0.05, Figure 2E).

**Figure 2.**
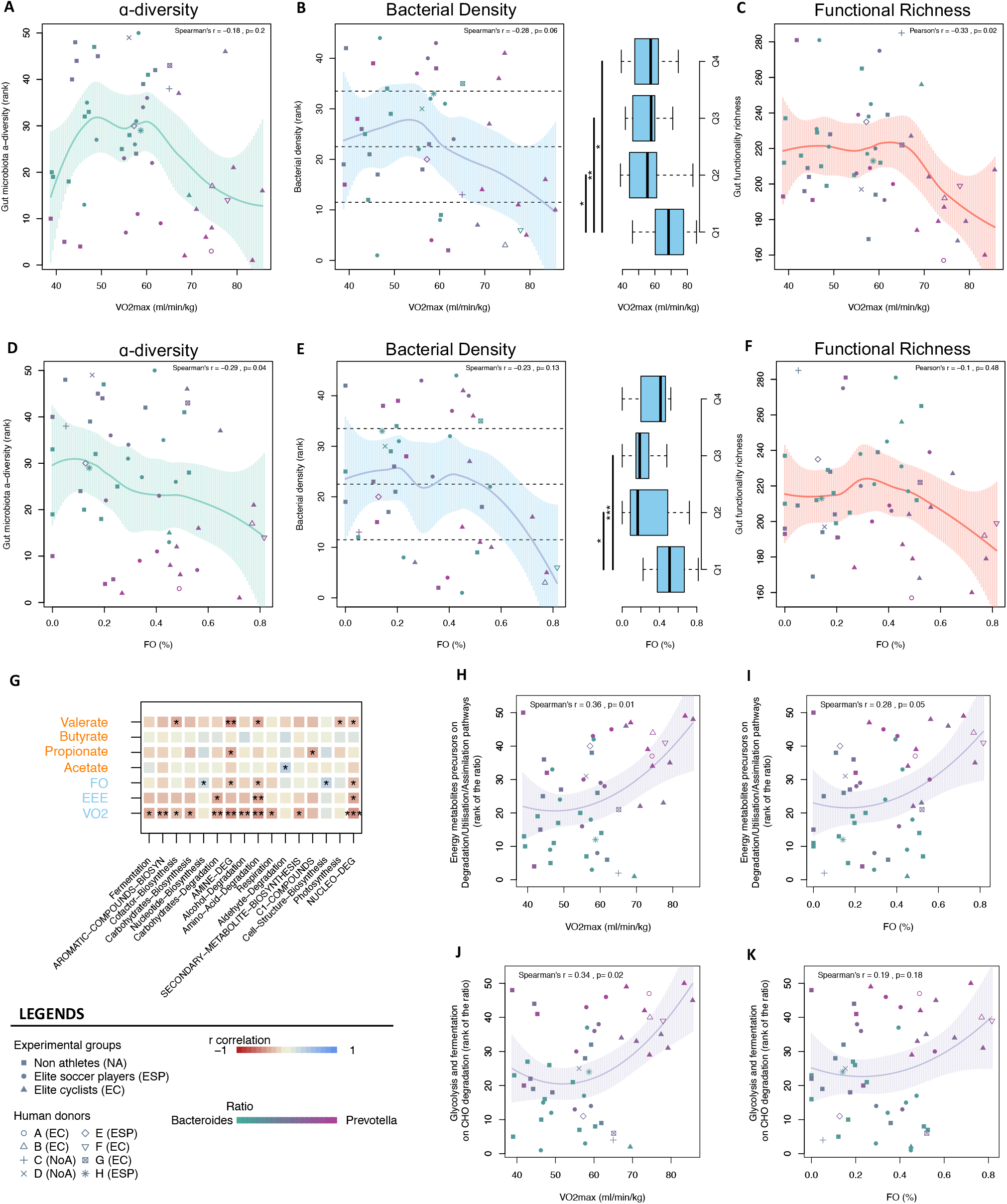
Metagenomic and metabolomic data support a selective pressure between gut microbiota and host energy metabolism. Local regression between 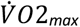 and gut microbiota *α*-diversity (**A**), bacterial density (**B**) and gut microbiota functionality abundance (**C**). Local regression between fat oxidation (FO %) and gut microbiota *α*-diversity (**D**), bacterial density (**E**) and gut microbiota functionality abundance (**F**). The local green, blue and red regressions correspond respectively to the *α*-diversity, bacterial density and functional abundance of the bacterial ecosystem within the EXOMIC cohort. The boxplots represent the repartition of VO_2max_ (**B**) and FO (%) (**E**) within each quantile of bacterial density. Quantiles are materialized by dashed lines on the local regression plots. (**G**) Correlation between gut microbiota functional families and fecal SFCAs, and exercise capacity parameters. Local regression between the ratio of Precursors of Energy metabolites on Degradation/Utilization/Assimilation pathways (gut microbiota functional superfamily) and 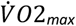 (**H**) or FO (**I**). Local regression between the ratio Glycolysis and Fermentation on CHO degradation pathways (gut microbiota functional superfamily) and 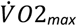 (**J**) or FO (**K**). For local regressions, the dark line represents the center of the local regression with the 95%-CI in dashed line.

To refine our analysis, we explored gut microbiota functionality at different levels according to the MetaCyc database. We observe that the ability of hosts to consume energy during maximal and submaximal exercises are inversely proportional to the richness of gut microbiota functionality (p<0.05, Figure 2C and S3C, respectively). Specifically, several classes of metabolic pathways, including fermentation and carbohydrates degradation, show a negative association with the hosts’ exercise capacity parameters, especially 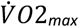 (Figure 2G). We then calculated the ratio between gut microbiota capacity to produce energy from available nutrients (i.e. abundance of pathways involved in the generation of precursor metabolites and energy), and its capacity to degrade substrates to serve as sources of nutrients to their own growth (i.e. Degradation/Utilization/Assimilation pathways abundance). Interestingly, this ratio is positively associated with the host’s ability to consume energy during maximal exercise (i.e. 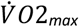, r = 0.36, p=0.01, Figure 2H). Notably, the ratio of glycolysis and fermentation pathways (i.e. pyruvate producers and fermenters) on carbohydrate degradation pathways (i.e. degradation, storage and utilization of CHO essential for the bacterial growth) is also positively associated with maximal energy consumption during exercise (r = 0.34, p=0.02, Figure 2H). Together, these findings support that hosts with high exercise capacities exhibit a reduction in gut functionality richness, but this reduction seems to be associated with a remodeling of the gut bacterial ecosystem and its metabolic pathways, favoring bacterial energy production from available nutrients over the production of energy from complex molecules (e.g. starch and fibers).

### Fecal microbiota transplantation (FMT) from high to very high exercise capacity human donors elicits distinct gut microbiota composition in transfected mice

To evaluate whether the atypical gut bacterial ecosystem observed in humans with very high exercise capacity could directly affect host’s energy metabolism and exercise capacity, we transfected gut microbiota from high to very high exercise capacity humans into ATB-pretreated mice (Figure 3A). We selected 8 physically active and non-sedentary donors exhibiting distinct exercise capacities (i.e. from high to very high 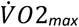, from low to very high FO) (Table S2). The fecal bacterial density, species engraftment rate, Bray-Curtis similarity between human donors and transfected mice post-FMT, as well as the donors gut microbiota relative abundance fractions reported post-FMT, are in accordance with current references to consider successful FMT engraftment in humans [38–40], thus supporting the effectiveness of FMT from human donors to transfected mice in our experiments (Figures S5A-E). The dissimilarity between mice reseeded with their own litter and transfected mice confirms such conclusions (Figures S5G-F). We next investigated the link between the parameters of donor exercise capacity with the gut microbiota compositions expressed in the transfected mice by using Canonical Analysis of PCo (CAP, Supplemental Material). We show that 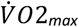, FO and EEE are associated with 17% of the variability of the gut microbiota compositions of transfected mice (p<0.01). The second axis of the canonical analysis (CAP2) refers to the mice gut microbiota compositions explained by fat oxidation (FO) and exercise energy expenditure (EEE) of the human donors (Figure 3B). Here, we specifically identified *Akkermensia municiphila, Parabacteroides merdae* and *Bacteroides vulgatus* among the bacteria of transfected mice well-associated with exercise capacity parameters (Figure 3B). Together, these findings highlight that distinct gut microbiota compositions are transferred from humans to transfected mice depending on the exercise capacity parameters of the donors.

**Figure 3.**
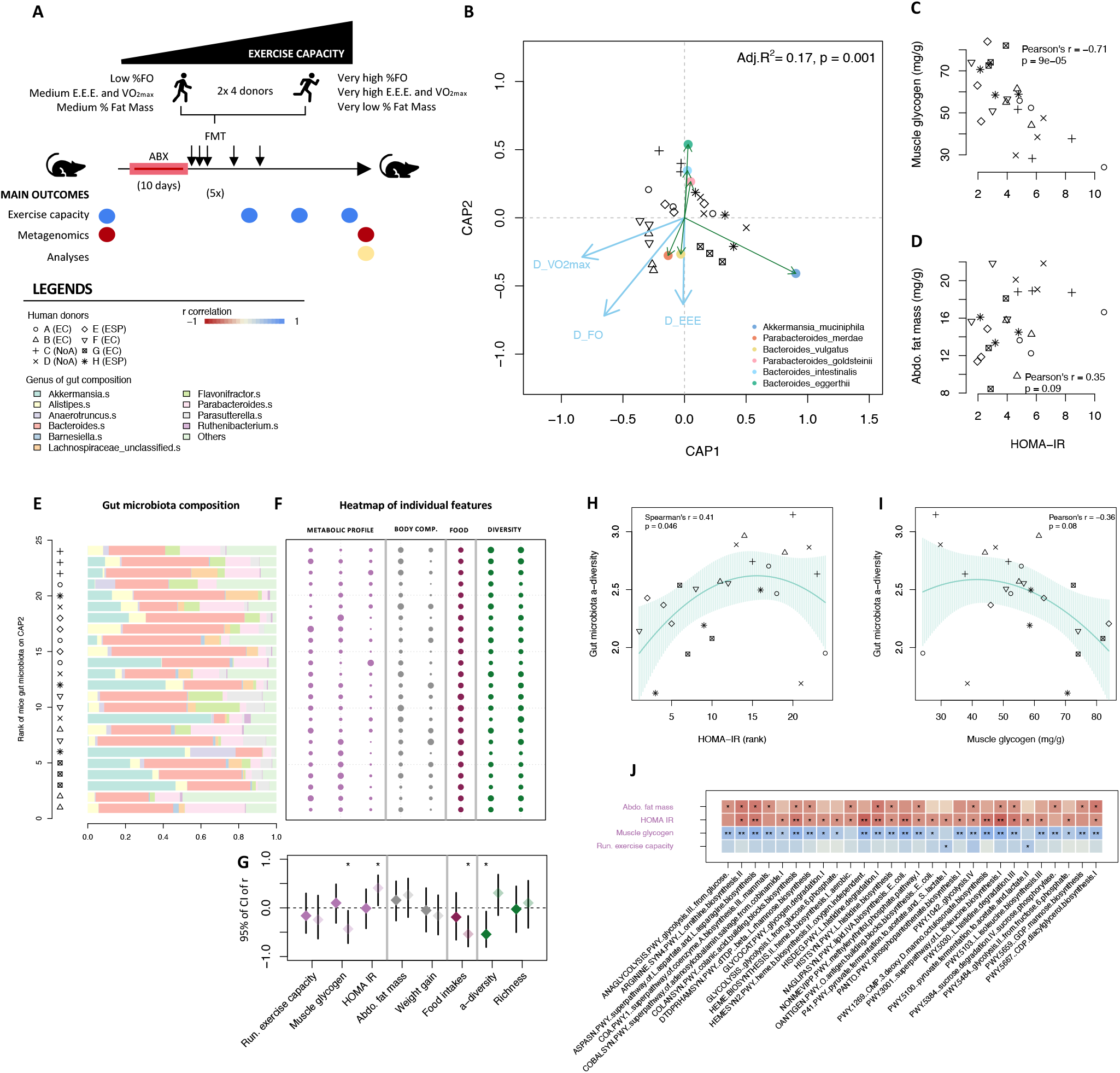
The gut bacterial ecosystem from human donors affects insulin sensitivity and muscle glycogen stores in transfected mice, depending on the donor’s exercise capacity. (**A**) Experimental design of FMT from human donors with heterogeneous exercise capacity into antibiotic-pretreated mice. (**B**) Canonical Analysis of Principal Coordinates (CAP) between the Principal Coordinates of the pairwise dissimilarity matrix (Bray Curtis) of mice’s gut microbiota compositions and the donors’ exercise capacity parameters (blue arrows). The points represent the most associated species of the second axis of the CAP. (**C**) Muscle glycogen content according to HOMA-IR. (**D**) Abdominal fat mass according to HOMA-IR. (**E**) Ranked mice from the CAP2 with the corresponding gut microbiota composition (1 line = 1 mice). (**F**) Heatmap of metabolic profile, body composition, food intake and gut microbiota composition’s diversities (1 line = 1 mice), the width of each point is normalized by the intra-variable maximum. (**G**) 95% confidence interval of the r correlation between each variable either with CAP1 (in dark colors) or with CAP2 (in light colors) determined in panel **B**. Local regression between gut microbiota *α*-diversity and HOMA-IR score (**H**) or muscle glycogen content in mice. (**J**) Correlations between gut microbiota functional families and metabolic parameters, exercise capacity and body composition in mice. Statistical significance of the correlation: **\*** p<0.05; **\*\*** p<0.01; **\*\*\*** p<0.001.

### The gut bacterial ecosystem from human donors affects insulin sensitivity and muscle glycogen stores in transfected mice, depending on the donor’s exercise capacity

Five weeks post-FMT, we assessed the running exercise capacity, HOMA-IR score and skeletal muscle glycogen content to characterize the exercise capacity and insulin sensitivity of each transfected mice (Table S3). We first observe that HOMA-IR score is positively correlated with relative abdominal fat mass, and negatively with skeletal muscle glycogen content (Figure 3C and 3D), indicating a heterogeneous continuum of metabolic profiles in the transfected mice.

On the Figures 3E and 3F, each line symbolizes variables characterizing a transfected mouse. The mice’s gut microbiota compositions are ordered depending on their positions in the second axis of the CAP (i.e. linked to the FO and the EEE of the donors). The Figure 3F represents the functional and metabolic variables (i.e. running exercise capacity, insulin sensitivity and body composition) associated to each gut microbiota composition. We next determined the 95%-CI of the r correlation between the second coordinate of the gut microbiota composition’s CAP and the physiological variables of the transfected mice. Here, we show that muscle glycogen content is inversely correlated with the second coordinate of the mice gut microbiota composition’s CAP (r = -0.49, p<0.05, Figure 3G), indicating that mice with gut microbiota compositions with negative coordinates in CAP2 exhibit a higher muscle glycogen level compared to those with positive coordinates. Since muscle glycogen content is directly associated with host insulin sensitivity [41], we logically observe that the HOMA-IR score is positively correlated with CAP2 (r = 0.41, p<0.05, Figure 3G), meaning that mice with gut microbiota compositions with negative coordinates in CAP2 exhibit better whole-body insulin sensitivity compared to those with positive coordinates. As previously described, *Akkermensia municiphila, Parabacteroides merdae* and *Bacteroides vulgatus* are the main bacteria characterizing the negative coordinate of the second axis of the CAP (Figure 3B). Our data are in line with previous studies where these bacteria were identified to exert metabolic benefits, especially reducing insulin resistance and fat mass gain in obese populations [42–44] and preventing atherosclerotic lesions in rodents [44,45]. In parallel to the results, we report positive correlations between HOMA-IR score and gut microbiota α-diversity in transfected mice (r = 0.41, p=0.046, Figure 3H), whereas muscle glycogen content is negatively correlated with gut microbiota α-diversity (r = -0.36, p=0.08, Figure 3I). These results suggest that the metabolic benefits of gut microbiota transplantation from human donors with very high exercise capacity do not necessarily depend on the gut microbiota α-diversity.

We finally explored the gut metabolic functionality of transfected mice and the relationship between functional pathways, exercise capacity, insulin sensitivity and muscle glycogen content. We report a positive correlation between pyruvate fermentation into acetate and lactate with running exercise capacity, while these pathways are negatively associated with HOMA-IR score (p<0.05, Figure 3J). Similarly, we observe a positive correlation between pathways of pyruvate synthesis (Glycolysis IV, Glycolysis from glucose) and muscle glycogen content (p<0.05, Figure 3J). Our results are consistent with previous studies reporting that high levels of the functional pathways involved in glycolysis (i.e. pyruvate synthesis) in the gut increase the availability of pyruvate [46], a key substrate for functional pathways of fermentation synthetizing SCFAs from pyruvate degradation [47,48]. In agreement with previous studies [49], all these findings highlight that transfection of gut microbiota associated with specific donor characteristics, here exercise capacity parameters, can significantly remodel gut microbiota composition and exert metabolic effects in transfected recipients.

## DISCUSSION

During the last decade, substantial observational and interventional studies have been conducted to characterize the impact of physical activity on the gut microbiota ecosystem in humans, providing contradictory results concerning the effects on gut microbiota composition’s α-diversity in men and women [36,50–52]. The lack of consideration for the impact of diet when evaluating the effects of exercise capacities on gut microbiota composition is probably the main factor that can explain such discrepancies [28,36,51]. In our prospective clinical study, we rigorously recruited participants with low exercise capacity by matching their food intakes to match qualitatively those of participants with very high exercise capacity. Our data highlight that, for populations that do not present extreme exercise capacities (very low or very high), the effects of host’s exercise capacity on the gut bacterial ecosystem appear to be very modest compared to those of food habits.

On the contrary, our results support that the gut bacterial ecosystem is remodeled in individuals with a very high ability to consume energy and preferentially use fat during submaximal exercise. In these athletes, we report a clear reduction of gut bacterial diversity (i.e. α-diversity, bacterial density and richness of functionality) with a gut bacterial ecosystem dominated by *Prevotella copri*. This bacterial ecosystem is associated to high fecal propionate levels, in accordance with the role of this SCFA on exercise capacity and fat oxidation [19,35]. We also report that the ratio between functional pathways characterizing the ability to produce energy from available nutrients, and its capacity to degrade substrates to serve as sources of nutrients is higher in humans with a very high ability to consume energy. Collectively, these findings suggest that the relationship between host energy metabolism and the gut bacterial ecosystem may be influenced by the availability of energy resources, which decreases when the host’s physical activity levels and exercise capacity are high. In this context, the gut bacterial ecosystem seems to specialize to cope with this situation, with some collateral implications, including a reduction in bacterial diversity and density.

The FMT have been recognized as a therapeutic tool to treat non-communicable diseases including obesity, diabetes, neuropsychiatric disorders or inflammatory bowel diseases [53]. One of the current issues is to determine which clinical or metabolic characteristics the donor must possess to observe clinical and/or metabolic effects in the recipient. Numerous studies have already assessed the impact of FMT from donors with distinct metabolic profiles (i.e. mice, brown bears or humans), reporting benefits on transfected mice concerning glucose tolerance, insulin sensitivity or lipid metabolism [49,54,55]. While exercise capacity is a clinical hallmark of health status [1,2], it is not yet considered a donor’s clinical parameter that could induce metabolic benefits in recipients. However, few recent studies have highlighted that FMT from exercise-trained donor mice improves the metabolic profile of the transfected mice compared to those from sedentary donor mice [56,57]. In the present study, we intend for the first time to evaluate the consequences of FMT from human donors with high to very high exercise capacity on both metabolic parameters and exercise capacity in recipient mice. When we transfected ATB-pretreated mice with fecal samples from our human cohort, we observe in the transfected mice that donors with very high exercise capacity promote an overexpression of *Akkermensia muciniphila, Parabacteroides merdae* and *Bacteroides vulgatus*, recognized to induce metabolic benefits for the host [42,43,45]. Interestingly, these bacterial ecosystems are associated with improvements in whole-body insulin sensitivity and in muscle glycogen stores, supporting metabolic benefits associated to the transfer of gut bacterial ecosystem from donors with very high exercise capacity. Finally, mice exhibiting greater insulin sensitivity and higher muscle glycogen stores display a reduction of *α*-diversity and a higher expression of SCFA producing-pathways, further supporting the hypothesis of a specialization of the gut bacterial ecosystem in favor of metabolite generation, but detrimental for bacterial growth and diversity.

In a therapeutic perspective, we selected human donors with high to very high 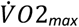 and normo-weight, constituting typical lean, healthy, very physically active donors. Despite gut microbiota remodeling and metabolic adaptations, the running exercise capacity of transfected mice remained unaffected. Importantly, these mice did not perform regular physical exercise during the 5 weeks of the FMT protocol, and the gut bacterial ecosystem they received from human donors was consequently not subjected to the high energy demands related to physical exercise. These results suggest that gut microbiota remodeling from donors with high exercise capacity alone is not sufficient to improve the exercise capacity of recipients. Overall, all these findings open promising research perspectives: 1) to improve the management of the gut microbiota ecosystem of elite athletes and patients performing adapted physical activity for therapeutic purposes, and 2) to personalize FMT in patients treated for non-communicable diseases by including exercise capacity in the clinical criteria for donor selection.

## Data Availability

All data produced are available online at GitHub https://github.com/DJrMartin/Gut-Microbiota-and-Physical-Activity.git

## DATA AND CODE AVAILIBILITY

Raw metagenomics sequencing data have been deposited into the NCBI under the accession number PRJNA1115089. In addition, the scripts and others data (metabolomics, food intakes and physiological data) used for the computational analyses described in this study are available at GitHub https://github.com/DJrMarhn/Gut-Microbiota-and-Physical-Activity.git. Any additional information required to reanalyze the data reported in this paper is available from the lead contact upon request.

### ACKNOWLEDGEMENTS

We thank all the participants and coaches from Stade Rennais FC and Sojasun Espoir for their committed efforts in participating in the present study. F.D. acknowledges fundings from the Brittany Council (N°21004090), the Société Française de Nutrition (SFN) and Nahibu. V.B. and F.D. acknowledge fundings from the University of Rennes. D.M, F.D and V.B acknowledge fundings from French National Research Agency within the framework of the PIA France 2030 programme for EUR DIGISPORT (ANR-18-EURE-0022) and CONTINUUM (ANR-21-ESRE-0030) projects.

## AUTHORS CONTRIBUTION

Conceptualization and methodology, D.M., M.B., A.C., V.B. and F.D.; data curation, D.M., M.B., M.H., L.O., R.D., M.H., E.L., R.B., A.G., P.G., M.C., F.D.; formal analysis and investigation, D.M., M.B., M.H., A.C., V.B. and F.D.; interpretation of data, D.M., M.B., V.B. and F.D.; writing – original draft, D.M.; writing – review & editing, V.B. and F.D.; funding acquisition, P.C., C.C., V.B. and F.D.; project administration, V.B. and F.D.; supervision, V.B. and F.D. All authors approved the final version of the manuscript.

