## Supplemental material for "Atypical gut microbial ecosystem from athletes with very high exercise capacity improves insulin sensitivity and muscle glycogen store in mice"

[1] Univ Rennes, M2S - UR 1274, F-35000 Rennes, France. [2] Univ Rennes, CNRS, IRMAR - UMR 6625, F-35000 Rennes, France. [3] Nahibu, Rennes, France. [4] ELS Consulting, Rennes, France. [5] Rennes Ortho Sport, Polyclinique Saint Laurent, Rennes, France. [6] Stade Rennais Football Club, Rennes, France. [7] CHU Pontchaillou, Rennes, France. [8] Institut du thorax, Nantes Université, CNRS, INSERM, Nantes, France. [9] UMS 016, UMS 3556, Nantes Université, Inserm, CNRS, Nantes, France. [10] IUT Vannes, University of South Brittany, Vannes, France

[#]: The last co-authors equally contributed to this work

Correspondence to: Frederic Derbré, PhD, Assistant professor. Laboratory "Movement, Sport and Health Sciences" – EA7470. University of Rennes 2 / ENS Rennes, 35170 Bruz, France. Mail:. Valerie Monbet, PhD, Full professor. IRMAR - UMR CNRS 6625. University of Rennes, 35000 Rennes, France. Mail:

**This file includes:**

**Supplementary materials and methods**

**Supplementary figures S1-S5**

**Supplementary tables S1-S5**

### Supplementary materials and methods

### EXPERIMENTAL MODEL AND SUBJECT DETAILS

#### Human clinical study

The EXOMIC study is a pilot clinical study that took place at the laboratory "Movement, Sport and health Sciences" (M2S), Rennes, France in 2022. The study was approved by the national Comité de protection des personnes Ouest IV Nantes (ID-RCB: 2021-A02496-35) and registered on ClinicalTrials.gov under NCT05220657. 50 male participants with low to very high exercise capacity were recruited from an elite cyclist team competing at the highest French national level (Elite cyclists, n=14), the academic structure of Stade Rennais FC, competing in the highest French national league (elite soccer players, n=15), and the student community of Rennes (Non-athletes, n=21). Non-athletes' participants were recruited from our local student community to fit with the food intakes of elite cyclists (limiting diet bias, more details in METHODS DETAILS OF STATISTICAL ANALYSIS). Inclusion criteria were being healthy, between 18 and 30 years of age, BMI (in $kg/m^2$) between 18 and 25, non-smoker and covered by medical insurance (more details on inclusion and exclusion criteria are available on ClinicalTrials.gov: NCT05220657). Each participant carried out three visits at the M2S lab: (1) a medical inclusion visit (2) a visit to perform an incremental maximal exercise test and (3) a last visit to perform a submaximal exercise test in fasted state. All participants gave their written informed consent. For each participant, the three visits were performed during a period of 15 days or less.

#### Mouse experiments

C57BL/6J male mice (8-week-old; Janvier Labs, France) were randomly divided into the following two experimental groups: control group (CTL, n = 6) versus mice transfected (n = 12) by heterogeneous human donors (n=4). Mice were maintained on a 12h/12-h dark-light cycle in a temperature-controlled room in individual cages with filter lids and had *ad libitum* access to food and water. The diet (MP-AL-S8189-S095, Janvier Labs) was standardized and identical for all groups. Except for running tests, mice were housed in individual standard cages without wheels, and thus were not subjected to daily exercise. Two independent campaigns of experiments were conducted to obtain a large sample size ($n_{\text{CTL mice}}$ = 12 ; $n_{\text{Transfected mice}}$ = 24 ; $n_{\text{Donors}}$ = 8). These animal experiments received approval from the Rennes committee on Ethics in Research (authorization APAFIS #39632-2022120109328170 v5) in accordance with the European directives (86/609/European Economic Community).

### METHOD DETAILS

#### Human clinical study

#### Medical inclusion visit (Visit n°1) and fecal samples collection

The first visit in the M2S lab consisted in a one-to-one interview including a medical exam with a physician. Each participant completed the food frequency questionnaire to estimate daily food and nutrient intakes (FFQ) [1]. Daily energy expenditure was also calculated on a weekly basis (MET-minutes per week) using the Global Physical Activity Questionnaire (GPAQ) [2]. Body Mass Index (BMI) was calculated as body mass in kilograms divided by height in square meters. Percentage body fat (%FM) was measured by skinfold measurement using Harpenden® forceps based on the 4-skinfolds method of Durnin and Womersley [3]. Body density (BD) was calculated using the same authors' equation based on skinfold measurements and the subject's age: BD = C - [M ($Log_{10}$ Σ (Skinfolds))], where C and M are coefficients varying according to sex and age [3]. The %FM was then calculated from BD using Siri's equation: %FM = 495/BD – 450 [4]. One fecal sample was collected for each participant during the 15 days following the medical inclusion and before the last visit to the laboratory. These samples were briefly stored by participants in a DNA blocker buffer for a maximum of one day before being frozen at -80°C in our partner laboratory (Nahibu, Rennes).

#### Incremental maximal exercise test (Visit n°2)

During the second visit to the laboratory, the participants performed an incremental test on ergocycle until reaching voluntary exhaustion to determine maximal oxygen uptake and the mechanical power developed for ventilatory thresholds 1 and 2. $\dot{V}O_2$ and $\dot{V}CO_2$ were continuously monitored during the test using an indirect calorimetry system (Ultima CardiO2, Medgraphics, United Kingdom). Heart rate and ECG signals were also continuously monitored during the test (12-lead Digital Holter Recorder). For soccer players and cyclists, the test began with a 3-minute warm-up at 150 watts, followed by 2 minutes at 175 watts. For non-athletes' participants, the test began with a 3 minutes warm-up at 100 watts, followed by 2 minutes at 150 watts. Then, the power was increased by 25 watts every 2 minutes until the subject could no longer maintain this power. Lactate blood concentrations were measured after each stage to determine lactate thresholds 1 and 2 using Lactate Pro 2 (Arkray, UK).

**Submaximal exercise test in fasting state (Visit n°3)**

During the last visit to the laboratory, the participants performed a submaximal test on ergocycle in a fasting state (between 7 and 10 a.m.) to determine carbohydrate and fat oxidation (FO). $\dot{V}O_2$ and $\dot{V}CO_2$ were continuously monitored during the test using an indirect calorimetry system (Ultima CardiO2, Medgraphics, United Kingdom). Heart rate and ECG signals were also continuously monitored during the test (12-lead Digital Holter Recorder). For all participants, the test began with a 4-minute warm-up at a power of 60 watts, followed by 10 minutes at the power developed at ventilatory threshold 1. A second stage, corresponding to 90% of the power developed at ventilatory threshold 2, was then performed during 10 minutes. The minute between 8 and 9 min of each stage was used to calculate absolute rates of fat and carbohydrate oxidation (g·min-1) using stoichiometric equations [5], based on the assumption that the excretion of urinary nitrogen was negligible. The proportion of fat oxidation in the total energy expenditure (FO, %) and the total energy expenditure during exercise (EEE, kcal/min) were calculated by summing the energy expenditure from both carbohydrate and fat oxidation.

**Mice experiments**

**Fecal Material Transfection (FMT) from humans to mice**

After 10 days of treatment with an oral cocktail of broad-spectrum antibiotics (1 mg/ml ampicillin, 5 mg/ml streptomycin, 1 mg/ml colistin, and 45 g/ml vancomycin in drinking water), mice were randomly assigned to FMT or control (CTL) groups. Drinking water containing antibiotics was changed every 60 hours during the 10-day treatment period. Mice selected for transfection (n=24) received 100 µg of fecal material by oral gavage every morning during 3 days, then once per week for the next two weeks. The fecal material was provided by the human donors from the EXOMIC cohort and preserved into a Maltodextrin/Trehalose (3:1) saline solution. In parallel, CTL mice (n= 12) underwent natural reseeding following the same 10-day antibiotic treatment. These CTL mice received two times soiled litter from their own litter, collected just before the antibiotic treatment. Fecal samples from FMT and CTL mice were individually collected before and after the antibiotic treatment, and 5 weeks post-FMT. 5 weeks post-FMT and overnight fasting, mice were weighed and glycemia was measured using Blood Glucose Monitoring System (Freestyle Papillon Vision, Abbott, France). Mice were then deeply anaesthetized with a ketamine-xylazine-butorphanol cocktail. Intracardiac blood was collected into dry tubes, and the mice were euthanized by cardiac exsanguination. Blood was then centrifuged (1500 g, 10 min, 4°C) for serum sampling. The liver, gastrocnemius muscles and abdominal fat mass were dissected, weighed and then either frozen in liquid nitrogen

**Running exercise capacity**

All mice performed a submaximal running test on a treadmill (Ugo Basile, Gemonio, Italy) before and after antibiotic treatment, and then each week during the next 2 weeks. Running exercise capacity was determined by the time until exhaustion from a test adapted from Okamoto et al. [6]. Briefly, the running speed started at 10 m/min for the first 10 min and then increased by 1 m/min every 14 min until reaching 24 m/min or exhaustion. Exhaustion was defined as the inability to maintain the normal running position and/or after five consecutive seconds in contact with the shock grid (0.2 mA) at the rear of the treadmill.

**Serum Parameters**

Glycemia was measured using Automated Beckman Coulter (Beckman Coulter, Brea, CA). Serum insulin concentrations were measured by enzyme-linked immunosorbent assay (ELISA) according to manufacturer's instructions (Millipore, St Louis, MO, USA). Insulin sensitivity was determined by calculating HOMA-IR according to the following formula: HOMA-IR = [fasting glucose (mmol/l)] × [fasting insulin (U/ml)] /22.5 [7].

**Muscle glycogen content**

The acid-hydrolysis method used for glycogen quantification was adapted from the protocols described by Adamo and Graham [8]. Frozen gastrocnemius muscle powder (20–30 mg) was placed in ice-cooled 6% perchloric acid and 1 M hydrochloric acid. Samples were then boiled at 100°C for 2 h, cooled on ice for 10 min, and centrifuged (1,300 revolutions/min for 5 min). Supernatant (12 µl) was removed from each sample, diluted in 200l of GOD-PAP solution (Biolabo, Maizy, France), and incubated in the dark at 37°C for 20 min to determine the glucose concentration. Absorbance was measured at 500 nm using a microplate reader (Varioskan™ LUX multimode microplate reader).

**Metagenomics data**

**Fecal DNA Extraction**

Total cellular DNA was extracted from 0.1 g of mice or human fecal material using the Quick-DNA Fecal/Soil Microbe Mini Prep Kit (ZymoResearch, CA, USA). Fecal samples were homogenized in the supplied cell suspension solution. A cell lysis/denaturing solution and 0.1-mm-diameter silica beads were then added, and the samples were mixed at maximum speed in a Beadbeater (Biospec, Bartlesville) for 1h at 4°C.

**Fecal DNA** quantification (Bacterial density)

The bacterial density (total amount of bacterial DNA) present in fecal samples of each mouse of human participant was evaluated by real-time qPCR targeting "all bacteria"16S rRNA genes using the universal primers F-bact1369 and R-prok1492. Experiments were performed using CFX Real-Time detection system (Bio-Rad Laboratories) with a final volume of 10 µL containing 5 µL of bacterial DNA (diluted at 1/2000), 0.2 µL of primers (10 µM), and 4.8 µL of SYBR® Green Super-mix (Bio-Rad Laboratories Inc., USA). All qPCR assays were performed in duplicate using the following cycling conditions: 50°C for 2 min and then 95°C for 2 min followed by 35 cycles of 95°C for 3 s and 60°C for 30 s. For the quantification, the E. coli DNA standard curve was generated by plotting the threshold cycles (Ct) versus bacterial quantity.

**Sequence pre-processing, taxonomic and functional profiling**

Fecal metagenomic shotgun sequences from both human and mice experiments underwent a pre-processing pipeline, where sequences were quality filtered using KneadData2 with default parameters [9]. MetaPhlAn3 was used for quantitative profiling the taxonomic composition of the microbial communities of all metagenomic samples [9]. HUMANn3 was used to profile pathway and gene family abundances [9].

**Metabolomic data**

Targeted metabolomic profiling of human fecal samples was performed by the CORSAIRE platform (Biogenouest, Nantes, France).

**Short Chain Fatty Acids**

Measurement of short-chain fatty acids (SCFAs) was performed as previously described with slight modifications [10]. A stock solution of SCFA metabolites was prepared and serially diluted to get ten calibration solutions. A working solution of internal standards was prepared in 0.15 M sodium hydroxide to get the following final concentrations: 75 mmol/L of $^{2}H_{3}$-acetate, 3.8 mmol/L of $^{2}H_{5}$-propionate, 2.5 mmol/L of $^{13}C$-butyrate, and 0.5 mmol/L of $^{2}H_{9}$-valerate. Fecal samples were weighed, homogenized at 0.1 g/mL in sodium hydroxide solution 0.15 M, and centrifuged 3 min at 7 000 g (4 °C). Samples (50 µL of fecal supernatants, serums or standard solutions) were dissolved in 200 µL of sodium hydroxide solution 0.15 M. Twenty microliters of the internal standard solution were added to samples and calibration solutions. After addition of the standards, each sample was acidified with 5 µL of hydroxide chloride 37% and then extracted with 1.7 mL of diethyl ether. Samples were stirred gently for 1h and then centrifuged 2 min (7000 g, 4 °C). The organic layers were transferred into 1.5 mL glass vials and SCFAs were derivatized with 20 µL of tert-butyldimethylsilyl imidazole. Samples were incubated for 30 min at 60 °C before analysis. Samples were finally analysed by gas chromatography–mass spectrometry (model 7890A-5975C; Agilent Technologies, Montpellier, France) using a 30 m x 0.25 mm x 0.25 µm capillary column (HP5-MS; Agilent Technologies). The temperature program started at 50 °C for 1 min, ramped to 90°C at 5 °C/min and then up to 300 °C at 70 °C/min. Selected ion monitoring mode was used to measure SCFA concentrations with ions at mass-to-charge ratio (m/z) 117 (acetate), m/z 120 ($^2H_3$-acetate), m/z 131 (propionate), m/z 136 ($^2H_5$-propionate), m/z 145 (butyrate and isobutyrate), m/z 146 ($^{13}C$-butyrate), m/z 159 (valerate), and m/z 168 ($^2H_9$-valerate). Chromatographic peak area ratios between unlabelled compounds and their respective internal standards constituted the detector responses. Standard solutions were used to plot calibration curves for quantification.

**Amino Acids**

Amino acid concentrations were determined in fecal samples by liquid chromatography-tandem mass spectrometry (LC-MS/MS) as described previously with slight modifications [11]. Fecal samples were weighed (50 mg) and dissolved in 500 µL of water. Fecal homogenates were centrifuged for 10 min (10 000 g, 4 °C) and supernatants were collected. Individual stock solutions (10 mmol/L) of unlabelled and labelled amino acids were prepared in 0.1 M HCl. A pool of unlabelled standard solutions was prepared and serially diluted in water to obtain seven standard solutions ranging from 0.01 to 100 µmol/L (equivalent to 0.1-100 nmol/g). A pool solution of labelled amino acids (10 µmol/L) was prepared in water. The standard solutions and fecal homogenate samples (20 µL) were then extracted with 100 µL of methanol and 50 µL of the labelled amino acid solution. The samples were mixed and centrifuged (10000 g, 4 °C) for 15 min to remove the precipitated proteins. The supernatants were collected and dried under a gentle stream of nitrogen (45 °C). The derivatization step was performed by dissolving the dried extract in 100 µL of a freshly prepared butanol solution containing 5% acetyl chloride and kept at 60 °C for 30 min. The solvent was then removed under a gentle stream of nitrogen (60 °C). The dried samples were dissolved in 100 µL of water containing 0.1% formic acid and 50 µmol/L tris (2-carboxyethyl) phosphine and injected into the LC-MS/MS system. Samples (10 µL) were injected onto an Acquity BEH-C18 column (1.7 µm; 2.1 × 100 mm, Waters Corporation) held at 60 °C, and compounds were separated with a linear gradient of mobile phase B (0.1% formic acid in methanol) in mobile phase A (0.1% formic acid in water) at a flow rate of 400 µL/min. Mobile phase B was kept constant at 1% for 0.5 min, linearly increased from 1% to 95% for 4.5 min, kept constant for 1 min, returned to the initial condition over 0.5 min, and kept constant for 1.5 min before the following injection. Target compounds were then detected by the mass spectrometer with the electrospray interface operating in the positive ion mode (capillary voltage, 3 kV; desolvation gas ($N_2$) flow, 650 L/h; desolvation gas temperature, 350 °C; source temperature, 120 °C). The multiple reaction monitoring mode was applied for MS/MS detection as detailed in the Table S4. Standard solutions were used to plot the calibration curves for quantification.

**Bile Acids**

Bile acids (BAs) were analysed in fecal samples using LC–MS/MS as previously described [12]. Fecal samples were weighed (50 mg) and dissolved in 500 µL of water. A pool of reference standard solutions including cholic acid (CA), chenodeoxycholic acid (CDCA), deoxycholic acid (DCA), ursodeoxycholic acid (UDCA), hyodeoxycholic acid (HDCA) and lithocholic acid (LCA) was prepared and serially diluted in water to obtain 7 standard solutions ranging from 0.1-100 µg/mL (equivalent to 0.1-100 µg/g). Ten microliters (10 µL) of $^2H_4$-CA at 100 µg/mL in 50% methanol were added to 500 µL of standard solutions and fecal samples. All samples were then mixed in an ultrasonic bath for 15 min. After complete homogenization, samples were acidified with 25 µL of 37% hydroxide chloride and then extracted with 1.2 mL of ethyl acetate (Biosolve, France). Samples were centrifuged for 10 min (5000 g, 4 °C). Supernatants were collected and dried under a gentle stream of nitrogen. Dried samples were finally dissolved in 600 µL of 50% acetonitrile and injected (10 µL) into the LC–MS/MS. LC–MS/MS analyses were performed using a Xevo® TQD mass spectrometer with an electrospray interface and an Acquity H-Class® UPLC™ device (Waters Corporation, Milford, MA, USA). Samples (5 µL) were injected into a CORTECS UPLC C18 column (1.6 µm; 2.1 × 100 mm, Waters Corporation) held at 60 °C, and compounds were separated with a linear gradient of mobile phase B (50% acetonitrile, 50% isopropanol, 0.1% formic acid, and 10 mM ammonium formate) in mobile phase A (5% acetonitrile, 0.1% formic acid, and 10 mM ammonium formate) at a flow rate of 400 µL/min. Mobile phase B was maintained at a constant volume of 10% for 1 min, linearly increased from 10% to 60% for 9 min, linearly increased from 60% to 95% for 1 min, held constant at 95% for 1 min, returned to the initial condition (10%) over 1 min, and held constant for 1 min before the following injection. Targeted compounds were then detected using the mass spectrometer with the electrospray interface operating in negative ion mode (capillary voltage, 2 kV; desolvation gas ($N_2$) flow rate and temperature, 1000 L/h and 400 °C, respectively; source temperature, 150 °C). Multiple reaction monitoring mode was applied for MS/MS detection as detailed in the Table S5. Standard solutions were used to plot the calibration curves for quantification.

### QUANTIFICATION AND STATISTICAL ANALYSIS

#### Non-athlete participants inclusion

We performed a pre-selection of the non-athlete participants based on their food habits. First, we estimated the nutrient and food intakes of the first 12 cyclists included in the EXOMIC cohort, recorded by a short-form food frequency questionnaire [13]. Each non-athlete participant was included if their macronutrient intakes were comprised into mean ± 2 standard variations of the macronutrient intakes measured in the cyclist groups. We also recorded the level of physical activity that each potential participant performed weekly. We aimed to recruit 10 participants under the WHO recommendations for physical activity levels, and 10 just above these recommendations (i.e. 600 METS/min/week). Out of the 102 non-athletes' volunteers we pre-selected, only 21 matched with these criteria and were included in the EXOMIC cohort.

#### Data normalization

For the microbiome data, we performed a Total Sum Scaling (TSS) within each metagenome. In this case, all the participants exhibit a sum of species equal to 1. We summed species at their phylogenetic genus level to 1. By using this new dataset, we were able to calculate the ratio between species belonging to the genus *Prevotella* and species belonging to the genus *Bacteroides*. For the metabolomic data, we performed a normalization of the fecal metabolite content based on the water content in their fecal material. Consequently, each volunteer exhibited metabolite content (mol) per g of dry faeces. For the nutrient intakes, we performed a total sum scaling within participants, except for the caloric intake, where we divided the total energetic intake by the participant's body mass (BM). We calculated caloric intake by multiplying CHO and protein intake by 4 and fat by 9.

#### Exploratory methods on bacterial ecosystem

To assess the intra-sample diversity of the bacterial ecosystem, we measured two indices: (A) the number of different species (richness) and (B) the evenness of the gut microbiota (Shannon index). These indices were computed using the *diversity* function from the *vegan* R package.

To assess the inter-sample diversity of the gut microbiota composition, we applied a Principal Coordinates Analysis [14,15] on the pairwise sample dissimilarity matrix of the gut microbiota compositions using Bray Curtis metrics. We presented the 20 strongest Spearman correlation between species and the two first Principal Coordinates. In other words, the 20 presented species are those that contribute the most to the Principal Coordinates Analysis of the gut microbiota. We also projected the donor's exercise capacity parameters as supplementary variable (Figure 3B). The coordinates were obtained by computing the Spearman's r correlation respectively with PCo1 and PCo2. The computation of pairwise sample dissimilarity matrix was performed using the *vegdist* function from the *vegan* R package, and its representation in the latent space (PCoA) was performed using the *pcoa* function from the *ape* R package.

**Interpretive methods on bacterial ecosystem**

Hierarchical Ascending Clustering (HAC) was used to cluster the participants according to their gut microbiota. The pairwise dissimilarity sample matrix essential for HAC is the Bray-Curtis metrics. The dendogram shows the merging sequence. The HAC was computed with the *hclust* function in R, using the *ward.D* agglomerative method.

Correlation describes the strength and direction of the relationship between two quantitative variables. In our results, we consistently presented two statistics: (a) the p value of the test where null hypothesis is no correlation, (b) the r correlation that provide the direction of the relation. The Pearson's r correlation was presented if the data respected homoscedasticity and normality hypothesis. When these conditions were not met, the Spearman's r correlation was used. Furthermore, variables not following a normal distribution are presented as rank statistics in the associated plot. The 95% confidence interval was computed by estimating the correlations for 1000 bootstrap samples.

Local linear regression was estimated to assess the variability and heterogeneity in relationships between two variables. Local linear regression is typically estimated by defining a neighbourhood around each data point and then fitting a linear model (such as linear regression) within that window to estimate the local relationship between the variables. This was performed using the *loess* function in R.

Canonical Analysis of Principal Coordinates (CAP) is the non-parametric form of the Redundancy Analysis (RDA) [16]. It is a multivariate statistical technique exploring the relationships between sets of variables. CAP measures how much environmental variables (i.e. diet, exercise capacity parameters, SCFAs) are linked to the structure and composition of the bacterial ecosystem. CAP is composed of two steps; in the first one, the PCoA of the species variables is performed; in the second step a canonical correlation analysis is computed that maximizes the correlation between the environmental variables and the axis of the PCoA of step 1. The results of CAP can be interpreted in terms of relationships between environmental variables and bacterial ecosystem. In the results, we obtained two statistics: (a) the adjusted explained variance ($R^2$) of Y (gut microbiota compositional data) by the set of variables X (environmental variables), and (b) the p value associated with the test where the null hypothesis is that X is linearly independent of Y. The used function was *capscale* provided by the *vegan* R package.

### Supplementary figures S1-S5

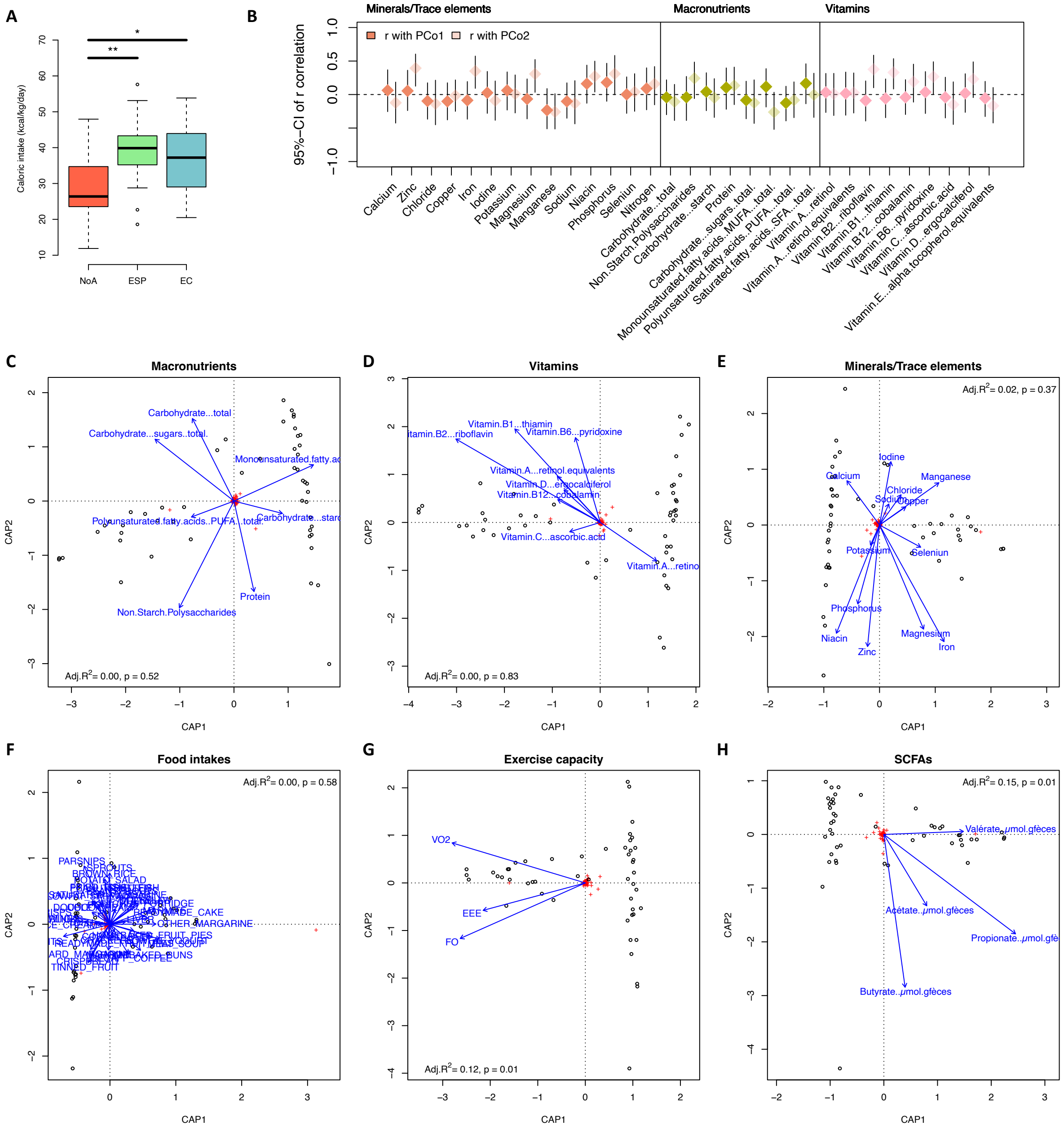


**Supplementary figure S1: Relationships between gut microbiota composition, food and nutrient intakes, exercise capacity and SCFAs.** (A) Caloric intakes calculated from FFQ in the three experimental groups of the EXOMIC cohort (*: p<0.05 and **: p<0.01, Wilcoxon rank sum tests) (B) 95% confidence interval for the r correlation between minerals, trace elements, macronutrients, vitamins calculated from FFQ with gut microbiota compositions (The dark and light points are related to PCo1 and PCo2, respectively). (C) Canonical Analyses of the Principal coordinates analysis (CAP) depending on macronutrient intakes, (D) vitamins, (E) trace elements and minerals, (F) food intakes, (G) exercise capacity parameters and (H) SCFAs. For each CAP, we represented two statistical parameters: (a) the adjusted explained variance ($R^2$) of gut microbiota compositions (Y) variability by the set of selected variables (X), and (b) the significance of the ANOVA test of the X explanation on Y (p value).

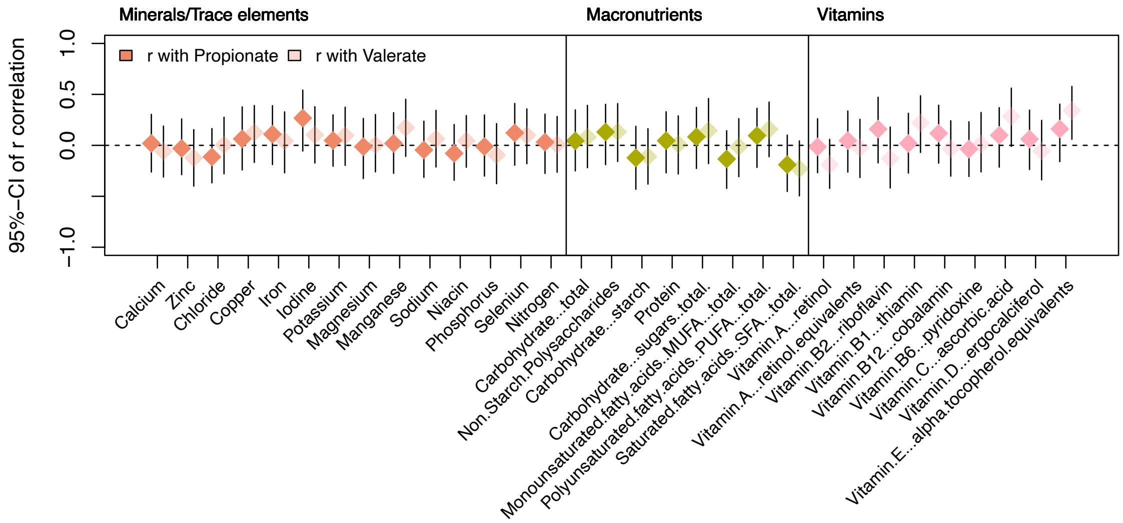


**Supplementary figure S2: Relationships between SCFAs and nutrient intakes.** 95% confidence interval for the r correlation between each nutrient intake calculated from FFQ with fecal propionate and valerate levels. The dark and light points are related to propionate and valerate, respectively

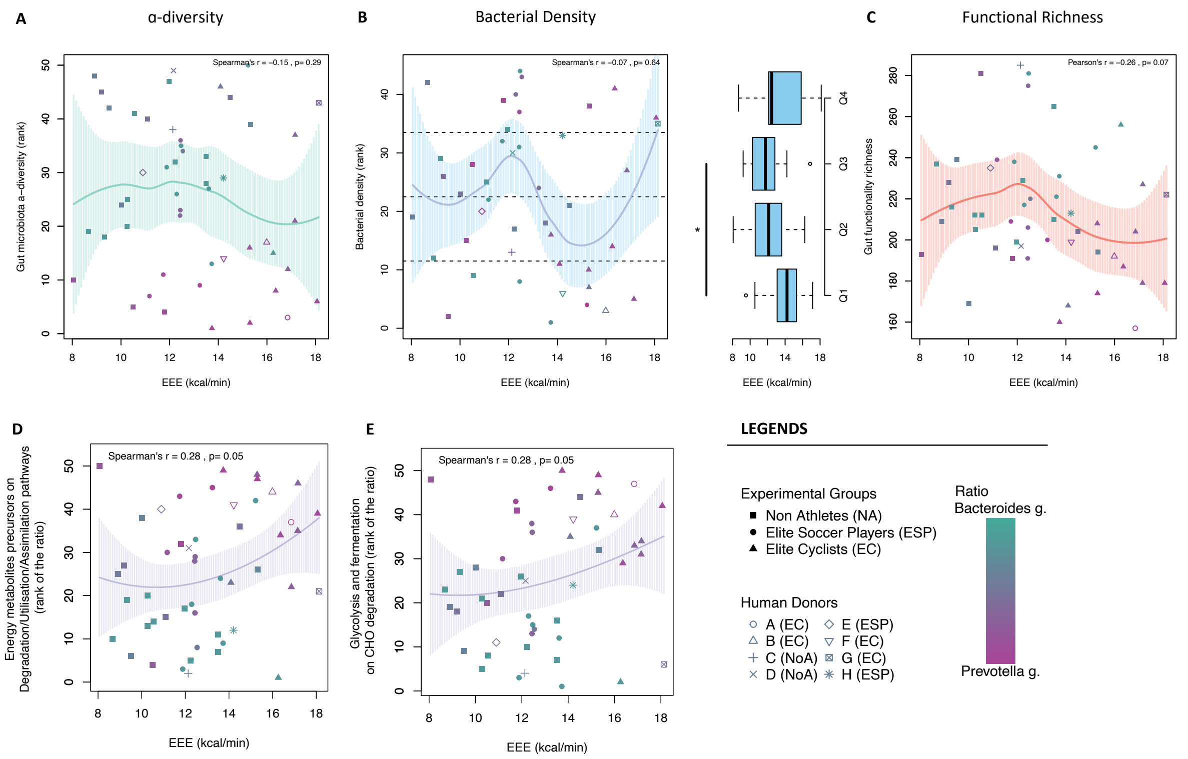


**Supplementary figure S3: Relationships between Exercise Energy Expenditure (EEE) and gut microbiota diversities.** (**A**) Local regression between EEE and gut microbiota composition α-diversity. (**B**) Local regression between EEE and ranked gut microbiota density. The boxplots represent the quantile distribution of gut bacterial density as a function of EEE. (**C**) Local regression between EEE with gut functionality richness. (**D**) Local regression between EEE and the ratio between Precursors of Energy metabolites pathways and Degradation/Utilization/Assimilation pathways. (**E**) Local regression between EEE and the ratio between Glycolysis and Fermentation pathways with CHO degradation pathways. For the local regressions, the dark line represents the center of the local regression with the 95%-CI in dashed lines.

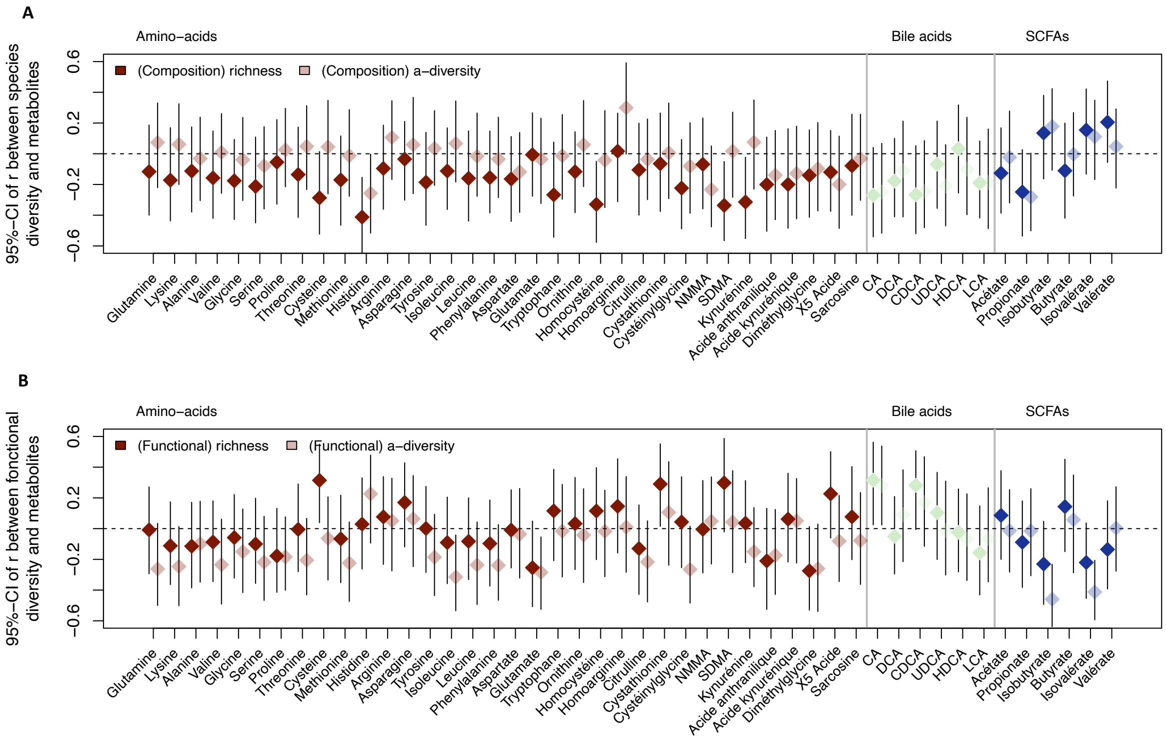


**Supplementary figure S4: Relationships between diversities of gut microbiota ecosystem and fecal metabolites.** (**A**) 95% confidence interval for the r correlation between each fecal metabolite content and species diversities. (**B**) 95% confidence interval for the r correlation between each fecal metabolite content and functional diversities. The dark and light points are related to the richness and the Shannon index, respectively.

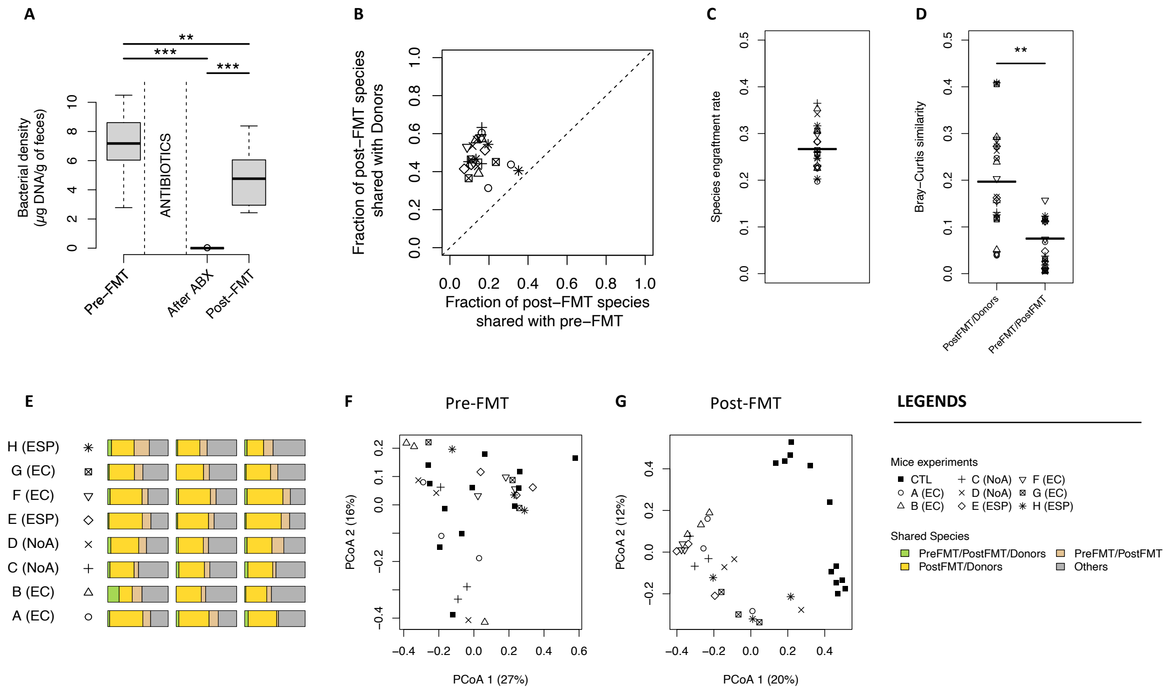


**Supplementary figure S5: Fecal Microbiota Transfection (FMT) leads to a unique donor gut microbiota community in transfected mice.** (**A**) Bacterial density (µg of DNA/g of feces) in transfected mice (n=15) at pre-FMT, after antibiotics (ABX) treatment and 5 weeks post-FMT (******: $p<0.01$; *******: $p<0.001$, Wilcoxon rank sum tests). (**B**) Fraction of post-FMT species shared with donors compared to the fraction of post-FMT species shared with pre-FMT. (**C**) Species engraftment rate (i.e. percentage of donor species present in the bacterial ecosystem of transfected mice (**, $p<0.01$, Wilcoxon rank sum test)). (**D**) Bray Curtis similarity between the gut microbiota compositions of donors and post-FMT mice, as well as between the gut microbiota compositions of pre-FMT with post-FMT mice. (**E**) Bar plot of the proportion of shared species in the transfected mice depending on their donors. (**F**) β-diversity of mice gut microbiota compositions from Bray Curtis pairwise dissimilarity matrix pre-FMT and (**G**) 5 weeks Post-FMT.

### Supplementary tables S1-S5

| CLINICAL FEATURES | Non-athletes (n = 21) | Elite soccer players (n = 15) | Elite cyclists (n = 14) |
|---|---|---|---|
| Age (years) | 22.81 ± 2.5 | 18.73 ± 0.7 [a] | 20.64 ± 2.0 [a,b] |
| Weight (kg) | 71.4 ± 8.3 | 74.4 ± 8.2 | 70.1 ± 7.1 |
| Height (m) | 1.79 ± 0.05 | 1.79 ± 0.07 | 1.79 ± 0.06 |
| IMC (kg/m$^2$) | 22.2 ± 2.2 | 23.1 ± 1.4 | 21.8 ± 1.5 [b] |
| Energy expenditure (MET.min/week) | 1032 ± 685 | 4724 ± 1612 [a] | 6039 ± 1995 [a, b] |
| Fat mass (%) | 15.1 ± 4.3 | 9.9 ± 1.5 [a] | 9.0 ± 2.9 [a] |
| Sedentary/Non-sedentary (n) | 18/3 | 2/13 | 4/10 |
| Active/inactive (n) | 13 / 8 | 15 / 0 | 14 / 0 |
| $\dot{V}O2_{max}$ (mL/min/kg) | 49.58 ± 8.20 | 56.21 ± 5.06 [a] | 74.34 ± 5.98 [a, b] |
| VT1 (% $\dot{V}O2_{max}$) | 53.1 ± 10.6 | 52.7 ± 5.2 | 57.8 ± 5.1 [a, b] |
| VT2 (% $\dot{V}O2_{max}$) | 74.8 ± 8.2 | 75.7 ± 5.7 | 72.8 ± 5.1 |
| Relative MPO (Watts/kg) | 3.76 ± 0.76 | 4.37 ± 0.43 [a] | 6.21 ± 0.46 [a, b] |
| Blood maximal lactate (mmol/L) | 14.2 ± 2.7 | 9.6 ± 3.1 [a] | 10.9 ± 3.0 [a] |
| Fat oxidation (kcal/min at VT1) | 1.97 ± 1.77 | 4.29 ± 1.57 [a] | 9.11 ± 2.41 [a, b] |
| Fat oxidation (kcal/min at VT2) | 1.71 ± 1.97 | 4.73 ± 1.95 [a] | 9.32 ± 3.51 [a, b] |
| CHO oxidation (kcal/min at VT1) | 9.15 ± 2.18 | 8.39 ± 1.73 | 7.97 ± 3.24 |
| CHO oxidation (kcal/min at VT2) | 12.6 ± 2.42 | 11.85 ± 3.12 | 8.53 ± 3.23 [a, b] |

**Supplementary Table S1: Clinical data of the participants included in the EXOMIC cohort.** Clinical features of non-athletes (NA, n = 21), elite soccer players (ESP, n = 15) and elite cyclists (EC, n = 14) are represented as mean ± SD. **[a]**: p<0.05 vs. NA; **[b]**: p<0.05 vs. ESP (Kruskal-Wallis followed by a Wilcoxon's post hoc test). Participants were considered sedentary if they spent more than 8 hours in a sitting position, and inactive if they performed less than 600 MET.min per week. CHO: carbohydrates, MPO: maximal power output, $\dot{V}O2_{max}$: Maximal Oxygen Uptake, VT1: Ventilatory Threshold 1, VT2: Ventilatory Threshold 2.

| Group | ID | Age range (years) | Weight (kg) | Height (m) | IMC (kg/m²) | $\dot{V}O2_{max}$ (ml/min/kg) | Fat ox. (kcal/min) | CHO ox. (kcal/min) | WEE (MET.min) | Fat mass (%) |
|---|---|---|---|---|---|---|---|---|---|---|
| **EC** | **A** | 21-25 | 79.0 | 1.83 | 23.5 | 74.3 | 8.23 | 8.62 | 4408 | 14.7 |
| **EC** | **B** | 18-20 | 61.1 | 1.72 | 20.0 | 74.5 | 12.30 | 3.69 | 3840 | 9.2 |
| **NoA** | **C** | 26-30 | 67.0 | 1.76 | 21.6 | 65.0 | 0.63 | 11.50 | 2040 | 13.0 |
| **NoA** | **D** | 20-25 | 77.9 | 1.85 | 22.8 | 56.1 | 1.86 | 10.30 | 3240 | 15.8 |
| **ESP** | **E** | 18-20 | 71.0 | 1.73 | 23.7 | 57.2 | 1.39 | 9.51 | 3600 | 12.0 |
| **EC** | **F** | 20-25 | 68.0 | 1.75 | 22.2 | 77.9 | 11.60 | 2.62 | 5120 | 8.0 |
| **EC** | **G** | 26-30 | 79.9 | 1.86 | 23.1 | 65.1 | 9.45 | 8.68 | 4800 | 8.8 |
| **ESP** | **H** | 18-20 | 70.2 | 1.70 | 24.3 | 58.7 | 2.01 | 12.20 | 9200 | 10.6 |

**Supplementary Table S2: Anthropometric and physiological characteristics of human donors for FMT.**

WEE: weekly energy expenditure, EC; elite cyclists, NoA: non-athletes, ESP: elite soccer players, $\dot{V}O2_{max}$: Maximal Oxygen Uptake.

| CLINICAL FEATURES | CTL (n = 12) | FMT (n = 24) |
|---|---|---|
| **Weight (g)** | 28.0 ± 1.2 | 27.5 ± 1.2 |
| **Food intake (g/g of weight per day)** | 0.160 ± 0.009 | 0.150 ± 0.011 ** |
| **Fat mass (mg/g)** | 13.81 ± 3.00 | 15.61 ± 3.59 |
| **Running exercise capacity (min)** | 171 ± 40 | 162 ± 42 |
| **Muscle glycogen Content (mg/g)** | 52.87 ± 24.01 | 54.89 ± 16.35 |
| **Serum insulin (ng/ml)** | 0.53 ± 0.29 | 0.47 ± 0.18 |
| **Glycemia (mg/dl)** | 162.3 ± 18.3 | 150.1 ± 23.8 |
| **HOMA-IR score** | 5.40 ± 3.52 | 4.43 ± 2.12 |

**Supplementary Table S3: Clinical and metabolic data from mice experiments.** Clinical features of control non-transfected mice (CTL, n=12) and transfected mice (FMT, n = 24) are represented as mean ± SD. Significant differences vs. CTL **: p<0.01 (Student's test)

**Supplementary Table S4: Multiple reaction monitoring parameters used for LC–MS/MS detection of amino acids.**

| **Amino acids** | **Precursor ion (m/z)** | **Fragment ion (m/z)** | **Cone (V)** | **Collision (eV)** |
|---|---|---|---|---|
| Acide Aspartique | 246.1 | 144.0 | 30 | 20 |
| $^{15}N$-Acide Aspartique | 247.1 | 145.0 | 30 | 20 |
| Acide Glutamique | 260.1 | 83.9 | 30 | 20 |
| $^{15}N$-Acide Glutamique | 261.1 | 84.9 | 30 | 20 |
| Alanine | 146.1 | 56.9 | 30 | 15 |
| $^{13}C$-Alanine | 147.1 | 56.9 | 30 | 15 |
| Arginine | 231.1 | 69.9 | 30 | 20 |
| $^{13}C$-Arginine | 232.1 | 69.9 | 30 | 20 |
| Asparagine | 189.1 | 73.9 | 30 | 20 |
| $^{15}N_2$-Asparagine | 191.1 | 74.9 | 30 | 20 |
| Cystéine | 178.1 | 75.9 | 30 | 15 |
| $d_3$-Cystéine | 181.1 | 78.9 | 30 | 15 |
| Glutamine | 203.1 | 83.9 | 30 | 20 |
| $^{15}N$-Glutamine | 204.1 | 84.9 | 30 | 20 |
| Glycine | 132.1 | 56.9 | 30 | 15 |
| $^{13}C$-Glycine | 133.1 | 56.9 | 30 | 15 |
| Histidine | 212.2 | 109.9 | 30 | 20 |
| $^{15}N_3$-Histidine | 215.2 | 112.9 | 30 | 20 |
| Isoleucine / Leucine | 188.2 | 85.9 | 30 | 15 |
| $^{15}N$-Leucine | 189.2 | 86.9 | 30 | 15 |
| Lysine | 203.1 | 83.9 | 30 | 20 |
| $^{13}C_6$-Lysine | 209.1 | 88.9 | 30 | 15 |
| Méthionine | 206.2 | 103.9 | 30 | 15 |
| $^{13}C$-$d_3$-Methionine | 210.2 | 107.9 | 30 | 15 |
| Phénylalanine | 222.2 | 120.2 | 30 | 20 |
| $d_5$-Phénylalanine | 227.2 | 125.2 | 30 | 20 |
| Proline | 172.1 | 69.9 | 30 | 20 |
| $^{13}C$-Proline | 173.1 | 70.9 | 30 | 20 |
| Sérine | 162.1 | 59.9 | 30 | 15 |
| $^{15}N$-Sérine | 163.1 | 60.9 | 30 | 15 |
| Thréonine | 176.1 | 73.9 | 30 | 15 |
| $^{15}N$-Thréonine | 177.1 | 74.9 | 30 | 15 |
| Tryptophane | 261.2 | 132 | 30 | 20 |
| $d_5$-Tryptophane | 266.2 | 137 | 30 | 20 |
| Tyrosine | 238.2 | 136 | 30 | 20 |
| $^{15}N$-Tyrosine | 239.2 | 137 | 30 | 20 |
| Valine | 174.1 | 71.9 | 30 | 20 |
| $^{13}C$-Valine | 175.1 | 71.9 | 30 | 20 |
| Anthranilic acid | 137.9 | 119.9 | 20 | 10 |
| $^{13}C_6$ –anthranilic acid | 144.0 | 126.0 | 20 | 10 |
| Kynurenic acid | 246.1 | 144 .0 | 30 | 25 |
| Quinolinic acid | 224.0 | 149.9 | 20 | 10 |
| Xanthurenic acid | 262.0 | 160.0 | 30 | 25 |
| $^{2}H_5$-kynurenic acid | 251.2 | 149.0 | 30 | 25 |

| | | | | |
|---|---|---|---|---|
| ADMA | 259.1 | 214.1 | 30 | 20 |
| NMMA | 245.1 | 69.9 | 30 | 20 |
| SDMA | 259.1 | 228.1 | 30 | 20 |
| $^{2}H_{7}$-ADMA | 266.2 | 221.1 | 30 | 20 |
| Citrulline | 232.1 | 112.9 | 30 | 20 |
| Homoarginine | 245.1 | 84.1 | 30 | 20 |
| Homocysteine | 192.1 | 89.9 | 30 | 15 |
| Cysteinylglycine | 235.0 | 76.0 | 20 | 20 |
| Cystathionine | 335.1 | 190.0 | 30 | 15 |
| $^{2}H_{7}$-citrulline | 237.2 | 118.0 | 30 | 20 |
| 5-HIAA | 248.0 | 146.0 | 30 | 15 |
| 3- Hydroxykynurenine | 281.2 | 152.0 | 30 | 15 |
| 5-Hydroxytryptophan | 277.2 | 260.1 | 20 | 15 |
| Kynurenine | 265.1 | 136.0 | 30 | 15 |
| $^{13}C_{3}.^{15}N$-hydroxykynurenine | 285.1 | 153.0 | 30 | 15 |
| 5-aminovaleric acid | 174.2 | 100.9 | 20 | 15 |
| Ornithine | 189.1 | 69.9 | 30 | 20 |
| $^{13}C_{5}$-ornithine | 194.1 | 73.9 | 30 | 20 |
| Sarcosine | 145.9 | 89.9 | 30 | 10 |
| Dimethylglycine | 160.0 | 103.9 | 20 | 10 |
| $^{2}H_{6}$-dimethylglycine | 166.0 | 110.0 | 20 | 10 |

**Supplementary Table S5: Multiple reaction monitoring parameters used for LC–MS/MS detection of bile acids.**

| Bile acids | Precursor ion (m/z) | Fragment ion (m/z) | Cone (V) | Collision (eV) |
|---|---|---|---|---|
| CA | 407.3 | 343.3 | 70 | 34 |
| DCA | 391.3 | 343.3 | 70 | 34 |
| CDCA, UDCA, HDCA | 391.3 | 391.3 | 70 | 15 |
| LCA | 375.3 | 375.3 | 70 | 15 |
| $d_4$-CA | 411.3 | 347.3 | 70 | 34 |

10 Cuffaro B, Assohoun ALW, Boutillier D, *et al.* Identification of New Potential Biotherapeutics from

Human Gut Microbiota-Derived Bacteria. *Microorganisms*. 2021;9:565. doi: 10.3390/microorganisms9030565

11 M W, M C, S R, *et al.* Nutritional biomarkers and heart failure requiring hospitalization in patients with type 2 diabetes: the SURDIAGENE cohort. *Cardiovasc Diabetol*. 2022;21:101–101. doi: 10.1186/s12933-022-01505-9

12 Brochard C, Bouguen G, Olivier R, *et al.* Altered epithelial barrier functions in the colon of patients with spina bifida. *Sci Rep*. 2022;12:7196. doi: 10.1038/s41598-022-11289-3

13 Cleghorn CL, Harrison RA, Ransley JK, *et al.* Can a dietary quality score derived from a short-form FFQ assess dietary quality in UK adult population surveys? *Public Health Nutr*. 2016;19:2915–23. doi: 10.1017/S1368980016001099

14 Borcard D, Gillet F, Legendre P. *Numerical Ecology with R*. Cham: Springer International Publishing 2018.

15 Xia Y, Sun J, Chen D-G. *Statistical Analysis of Microbiome Data with R*. Singapore: Springer 2018.

16 Anderson MJ, Willis TJ. Canonical Analysis of Principal Coordinates: A Useful Method of Constrained Ordination for Ecology. *Ecology*. 2003;84:511–25. doi: 10.1890/0012-9658(2003)084[0511:CAOPCA]2.0.CO;2
